# Direct and Indirect Genetic Effects on Early Neurodevelopmental Traits

**DOI:** 10.1101/2024.01.24.24301734

**Authors:** Laura Hegemann, Espen Eilertsen, Johanne Hagen Pettersen, Elizabeth C. Corfield, Rosa Cheesman, Leonard Frach, Ludvig Daae Bjørndal, Helga Ask, Beate St Pourcain, Alexandra Havdahl, Laurie J. Hannigan

**Affiliations:** Department of Psychology, University of Oslo, Oslo, Norway; Nic Waals Institute, Lovisenberg Diaconal Hospital, Oslo, Norway; PsychGen Center for Genetic Epidemiology and Mental Health, Norwegian Institute of Public Health, Oslo, Norway; PROMENTA Research Center, Department of Psychology, University of Oslo, Oslo Norway; Department of Child Health and Development, Norwegian Institute of Public Health, Oslo, Norway; Department of Clinical, Educational and Health Psychology, Division of Psychology and Language Sciences, University College London, London, UK; MRC Integrative Epidemiology Unit (IEU), University of Bristol, Bristol, United Kingdom; Language and Genetics Department, Max Planck Institute for Psycholinguistics, Nijmegen, the Netherlands; Donders Institute for Brain, Cognition and Behaviour, Radboud University, Nijmegen, the Netherlands

## Abstract

**Background:** Neurodevelopmental conditions are highly heritable and frequently co-occur within individuals. Recent studies have shown that SNP heritability estimates can be confounded by genetic effects mediated via the environment (indirect genetic effects). However, the relative importance of direct *versus* indirect genetic effects on early manifestations of neurodevelopmental conditions is unknown.

**Methods:** The sample included up to 24,692 parent-offspring trios from the Norwegian MoBa cohort. We use Trio-GCTA to estimate latent direct and indirect genetic effects on mother-reported neurodevelopmental traits at age three years (restricted and repetitive behaviors and interests, attention, hyperactivity, language, social, and motor development). Further, we investigate to what extent direct and indirect effects are attributable to common genetic variants associated with autism, ADHD, developmental dyslexia, educational attainment, and cognitive ability using polygenic scores (PGS) in regression modeling.

**Results:** We find evidence for contributions of direct and indirect latent common genetic effects contributing to attention (direct: explaining 4.8% of variance, indirect: 6.7%) hyperactivity (direct: 1.3%, indirect: 9.6%), and restricted and repetitive behaviors (direct effects: 0.8%, indirect effects: 7.3%). Variation in social and communication, language, and motor development was best explained by direct effects (5.1-5.7%). Direct genetic effects on attention were captured by PGS for ADHD, autism, educational attainment, and cognitive ability, whereas direct genetic effects on language development were captured by cognitive ability and autism PGS. Indirect genetic effects were primarily captured by educational attainment and/or cognitive ability PGS across all outcomes.

**Conclusions:** Results were consistent with differential contributions to neurodevelopmental traits in early childhood by direct and indirect genetic effects. Indirect effects were particularly important for hyperactivity and restricted and repetitive behaviors and interests, and may be linked to genetic variation associated with cognition and educational attainment. Within-family approaches are important for disentangling genetic processes that influence early neurodevelopmental traits, even when identifiable associations are small.

## Background

Neurodevelopmental conditions, such as autism and attention-deficit hyperactivity disorder (ADHD), are highly heritable^[1,2]^ and have the earliest age-of-onset of neuropsychiatric conditions.^[3]^ Prior to the ages at which these conditions are typically diagnosed, numerous early indicators have been robustly associated with later neurodevelopmental diagnoses. These include early childhood (< 3 years of age) behavioral traits^[4]^, infant developmental milestones^[5]^, and even intrauterine^[6]^ and gestational^[7]^ factors. In many cases, these early indicators have been linked to the genetic liabilities that underpin neurodevelopmental conditions.^[8–11]^ This is consistent with both the high heritability of these conditions and more than a decade of evidence indicating that the effects of most common genetic variants are diffuse across the neuropsychiatric phenome.^[12]^

This pattern of evidence is consistent with a relatively linear early etiology of neurodevelopmental disorders: strong genetic liabilities directly influence individuals’ gestation, early developmental milestones and behavioral traits, before ultimately manifesting as neurodevelopmental conditions when children are old enough for diagnoses to be made. In reality, causal structures underlying the development of complex human behaviors and the neuropsychiatric conditions comprising them are rarely so simple. In large part, this is because apparently distinct sources of liability are highly confounded, even from the earliest moments of life. The sharing of genetic material between parents and children means that even the earliest environments to which infants are exposed are correlated with their genotypes^[13,14]^. Additionally, due to the correlation between parental and child genotypes, biases arising from assortative mating, selection processes, population stratification, and – if relying on parental reported data – rater effects, can impact observed associations between a child’s genotype and potential early indicators^[15,16]^. This has implications not only for any assumptions we might make about the role of early environments for later conditions, but also for understanding their heritability.

Effects of an individual’s own genotype on their own behavior can be considered *direct genetic effects*. Such effects are commonly assumed to be central contributors to the heritability of any trait. However, another key source of heritable variation are *indirect genetic effects* – where variation in a trait in one individual is caused by the genotype of another individual^[17,18]^. In related individuals, although mediated via environmental processes, such effects appear linked to an individual’s own genotype. This is because they reflect heritable behaviors in other people who are genetically similar and often in close proximity (usually, though not always, parents or siblings). In the case of indirect effects of parents genotypes on their child’s phenotypes, these are sometimes referred to as “genetic nurture” effects.^[19]^ Of all the factors outside of direct effects that can influence estimates of the heritability^[20]^, indirect genetic effects are particularly interesting to isolate, because they represent – at least theoretically – environmental processes that can be detected without recourse to potentially unreliable or invalid observational environmental measures. Instead, genotype data from relatives can supplement genetic analyses of individuals to allow for disentangling direct and indirect genetic effects on phenotypes.

In recent years, researchers have increasingly begun using within-family approaches to disentangle direct and indirect genetic effects on neurodevelopmental traits and conditions. ^[16]^ The most straightforward approach, which allows direct genetic effects to be estimated without confounding by indirect effects and other structural confounders is to compare variant-trait associations within and between families. This can be done as a within-family genome-wide association study (GWAS), the largest example of which to date showed evidence that common variant heritability estimates for neurodevelopment-linked traits such as educational attainment and cognition may be reduced by as much as 50% when restricted only to direct genetic effects ^[21]^. Within-sibship polygenic score (PGS) analyses can address similar questions at a more granular level. In one such study in a UK-based twin sample, the prediction, by ADHD polygenic liability, of ADHD traits at age 12 was not significantly attenuated within-families (suggesting limited indirect genetic effects), although the prediction of educational performance was substantially (>50%) reduced. ^[22]^ Analogous approaches involve comparing the observed and expected transmission of polygenic liabilities among individuals diagnosed with neurodevelopmental conditions – where *over* transmission relative to expectations set by averaging parental genetic liabilities is evidence of direct effects. This approach has been used to confirm that direct genetic effects are at least partly responsible for the positive association between autism diagnoses and children’s PGS for autism, schizophrenia, and educational attainment. ^[23]^ A recent application of this method also found evidence of direct genetic effects on ADHD diagnoses captured in over-transmission of PGS for ADHD, and under-transmission of PGS for obsessive-compulsive disorder and cognitive ability. ^[24]^

Beyond seeking to estimate direct genetic effects free of confounding by indirect effects, it is also possible to proactively seek *evidence* of indirect genetic effects. Martin and colleagues ^[24]^ did this by comparing ADHD genetic liabilities in parental alleles not transmitted to their child to those among population controls. In contrast to the enrichment for ADHD PGS found in the transmitted parental alleles, they did not find evidence of elevated genetic liabilities for ADHD among non-transmitted parental alleles, which would have indicated a pathway for indirect genetic effects.^[24]^ Other approaches seek to estimate direct and indirect genetic effects in parallel. Among 8-year-old children in the Norwegian Mother, Father, and Child Cohort study (MoBa), two studies using variance decomposition methods – which seek to subdivide all variation in a trait that is linked to common genetic variants into either direct or indirect effects components – have found evidence consistent with modest (explaining 8-16% variance) indirect genetic effects on ADHD traits. ^[25,26]^ In both studies, *direct* genetic effects were estimated at around twice the magnitude of indirect effects. An alternative approach using PGS of family trios in the same dataset, found that only maternal genetic liability for neuroticism was a robust source of indirect genetic effects on 8-year-olds ADHD traits; whereas children’s genetic liability to ADHD, smoking, educational attainment, and cognitive ability were all potential sources of direct genetic effects. ^[27]^

Variance decomposition-based analyses estimate genome-wide latent direct and indirect genetic effects, while trio-PGS analyses seek to locate the signal of these effects within specific genetic liabilities. As such, these approaches have the potential to make distinct and complementary contributions in etiological investigations as they have differing underlying assumptions and limitations. Here, since little is known about the relative importance of direct and indirect genetic effects on early life neurodevelopmental traits, we apply both approaches in parallel to data from 3-year-old children in the MoBa cohort, addressing two main questions. Firstly, to what extent are genetic effects on early life neurodevelopmental traits likely to be direct *versus* indirect? Secondly, can these effects be attributed to established polygenic liabilities for neurodevelopmental conditions and related traits?

## Methods

### Measures and sample

#### Sample

The Norwegian Mother, Father and Child Cohort Study (MoBa) is a population-based pregnancy cohort study conducted by the Norwegian Institute of Public Health. ^[28,29]^ Participants were recruited from all over Norway from 1999-2008. The women consented to participation in 41% of the pregnancies. Blood samples were obtained from both parents during pregnancy and from mothers and children (umbilical cord) at birth.^[30]^ The cohort includes approximately 114,500 children, 95,200 mothers and 75,200 fathers. The current study is based on version 12 of the quality-assured data files released for research in January 2019. The establishment of MoBa and initial data collection was based on a license from the Norwegian Data Protection Agency and approval from The Regional Committees for Medical and Health Research Ethics. The MoBa cohort is currently regulated by the Norwegian Health Registry Act. The current study was approved by The Regional Committees for Medical and Health Research Ethics (2016/1702).

The present study was conducted on a subset of the cohort of complete genotyped family trios (N = 24,692) who had information available from the 3-year questionnaire. For more information on genotyping of the MoBa sample and for the family-based quality control pipeline used to prepare these data for analysis, see Corfield et al. ^[31]^. The sample for the Trio-GCTA analyses was further restricted to unrelated trios (N = 16,565). The children were on average 3.1 years old when mothers completed the questionnaire. The sample had a 1.05:1 male to female ratio.

#### Measures for neurodevelopmental traits

The *Social Communication Questionnaire* (SCQ) was used as a measure of social communication traits as well as repetitive and restrictive behaviors and interests (RRBI). The questionnaire was split into subscales of these two domains based on DSM-5 diagnostic criteria for autism (including 25 and 12 items respectively). Six items from the *Child Behavior Checklist* (CBCL) were split (3 items each) into two subscales to measure attention and hyperactivity. Motor development was measured using the *Ages and Stages Questionnaire* (ASQ). The MoBa questionnaire included two items from both the fine and gross motor subscales, which were combined to form an overall measure of motor development. Six items covering receptive and expressive language development (three items each) from the ASQ communication subscale were combined as a measure of language development. The exact items going into the scales can be found in the supplementary methods. Only overall motor and language scales (not subscales for fine/gross motor or receptive/expressive language) could be used in analyses that required continuous measures due to excessively skewed distributions. In sensitivity analyses, the fine motor and expressive language development subscales were treated as ordinal while receptive language and gross motor development were dichotomized so that all values greater than 0 were coded as 1 and interpreted as no versus any reported difficulties in these areas.

Scores for each measure were not calculated for individuals with greater than 50% missingness and these individuals were removed from the sample. For remaining individuals, the sum score was divided by the number of items answered and multiplied by the total number of items in the total score. This value rounded to the nearest whole number was presented as the total/subdomain scores. A square root transformation was performed on the measures of social communication, language, and motor development due to moderate right skewness and all measures were standardized to have a mean of 0 and standard deviation of 1. Items were reverse coded when necessary, so that for all scores higher values reflected the presence of more neurodevelopmental divergence.

#### Polygenic scores

Polygenic scores (PGS) were estimated with the software PRSice2 ^[32]^ based on summary statistics from the most recent GWAS for ADHD ^[33]^, autism ^[34]^ and dyslexia ^[35]^. These conditions were included as they are neurodevelopmental conditions with well powered GWAS. The PGS for dyslexia was created using the publicly available top 10,000 SNPs. In addition to including PGS for these neurodevelopmental conditions, PGS for educational attainment ^[36]^ and cognitive ability ^[37]^ were also estimated as they are related traits to neurodevelopment. The educational attainment PGS was constructed from summary statistics excluding 23andMe and MoBa participants. There is no direct overlap of MoBa participants and the other GWAS used. PGS for each of the five traits were derived based on creating scores at 11 different p-value thresholds between 5 × 10^-8^ and 1, then extracting the first principal component of these 11 scores for use as the PGS for that trait in the subsequent analyses. This approach controls for type one error rate arising from optimization of pruning and thresholding while still maintaining prediction performance ^[38]^. PGS were regressed on the first 10 genomic principal components (PCs) and genotype batch.

### Analyses

#### Trio-GCTA

To estimate the latent direct and indirect common genetic effects underlying early neurodevelopmental traits we use the Trio-GCTA ^[39]^ method. Trio-GCTA, an extension of earlier GCTA ^[40,41]^ methods, was developed to disentangle direct and indirect genetic effects. This is done by quantifying the additive genetic variance effects on an outcome from all measured single nucleotide polymorphisms (SNP) of all members of parent-offspring trios simultaneously. Indirect and direct genetic effects are partitioned using a linear mixed model, estimating variance components for all three members of the trios. In the current study, the child is the focal individual in the trio (i.e. the “owner” of the phenotype), so the estimated variance of the child’s genetic effects is interpreted as direct genetic effects. The estimated variances of the mother and father’s genetic effects are then interpreted as maternal and paternal indirect effects, respectively. In a “full” model, this includes genetics effects of the child (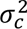; direct effect) as well as the genetic effects of the mother (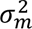; indirect effect) and father (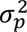; indirect effect). Additionally, the covariances between the child and mother (*σ_om_*), child and farther (*σ_op_*), father and mother (*σ_mp_*) genetic effects, as well as the residual variance (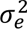), are estimated. The total variance decomposition of the phenotype is shown in Equation 1. While typically covariances would contribute to the total variance by a factor of two, for this model, we do not include the covariance between the father and mother in the calculation of the total variance. We do this under the assumption that parents are unrelated and have an expected genetic relatedness of 0. Additionally, since parents/children have a genetic relatedness of 0.5 we assume that these covariances contribute to the total variance by a factor of one instead of two.

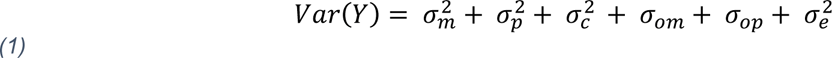

We next compared this “full” model to a nested model omitting the covariances between child/parent and parents’ genetic effects (“No covariance”). This “no covariance” model, in turn was compared to a nested model with only direct effects (“direct only”), which was subsequently compared to a “null” model including no genetic effects at all (Table 1). Model fit was assessed using Akaike’s Information Criteria (AIC). Additionally, Bayesian Information Criteria (BIC) was reported. When indices disagreed on the best fitting model, AIC was used. We chose AIC given that in the condition when the true model is not present in the candidate models, AIC is efficient (particularly when the true model is complex and/or contains small effects) at selecting the model closest to the true model while the BIC is not ^[42]^. Finally, a likelihood ratio test was preformed comparing the nested models for each outcome providing additional insight in the statistical significance of removing parameters from the model. P values from these tests were corrected for six tests (based on six outcomes) using false discovery method (FDR). Estimates of variance components were interpreted from the best fitting model but all results are presented given potential discrepancies in model selection.

**Table 1:** Variance decomposition of the four models tested in the Trio-GCTA analyses.

| Model | Variance decomposition |
| --- | --- |
| Full | $\sigma_c^2 + \sigma_p^2 + \sigma_m^2 + \sigma_{om} + \sigma_{op} + \sigma_e^2$ |
| No Covariance | $\sigma_c^2 + \sigma_p^2 + \sigma_m^2 + \sigma_e^2$ |
| Direct Only | $\sigma_c^2 + \sigma_e^2$ |
| Null | $\sigma_e^2$ |

The variance explained by genetic effects are estimated using 3 subsets (mothers, children, fathers) of a genomic relatedness matrix of the full sample ^[39]^. A threshold of 0.1 was applied to the matrix removing any individuals that had genetic correlations above this, ignoring parent/child relationships. This left 16,518 – 16,565 trios, depending on the outcome. Selection of individuals was done using the “bottom up” algorithm in OpenMendel ^[43]^. This was done to limit potential confounding due to closely related individuals^[20]^. The four nested models were run in Trio-GCTA for the six continuous neurodevelopmental trait measures: attention, hyperactivity, repetitive and restricted behaviors and interests, social communication development, motor, and language development. All models included fixed effects for child’s sex, genotyping batch, and first 10 genomic PCs.

#### Trio PGS regression modeling

Regression modeling was used to investigate direct and indirect effects attributable to PGS of neurodevelopmental conditions and related traits on measures of early neurodevelopment. The six continuous neurodevelopmental trait measures were used as outcomes in separate linear regression models. These measures were regressed onto the five PGS traits in independent models. In total this resulted in 30 models (6 continuous outcomes, each having 5 separate models for the 5 PGS traits) including the mother, father, and child PGS as predictors in each model. Jointly considering the effects of all family members in the trios allows for disentangling direct (transmitted) genetic effects from indirect genetic effects ^[44]^. Effect estimates from these models were interpreted as direct (*β*_1_; residual effect of the child’s PGS adjusting for parent’s PGS) and indirect effects (*β*_2_ and *β*_3_; residual effect of the mother and father’s PGS adjusting for the child’s PGS) of the PGS trait on the outcome. Sensitivity analyses looking into the subdomains of motor and language development were also run. Logistic regressions were run for binary versions of the gross and receptive language outcomes. Ordinal regression models were used for fine motor skills and expressive language development. This resulted in an additional 20 models.

A clustered sandwich estimator ^[45,46]^ was used to estimate standard errors for all models, with individuals clustered on maternal ID to account for nuclear family structures in the MoBa. Sex and age at time of questionnaires return date were included as covariates in the models, meaning that the standard model for a given outcome was as follows:

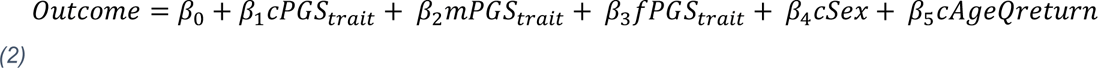

Individuals with reported age greater than 4 years were removed from the sample. P-values were adjusted for multiple testing across all regression analyses. These were calculated using the FDR method, correcting for 50 tests (10 outcomes x 5 PGS-traits).

In addition to models looking at effects of the PGS on neurodevelopmental traits individually (“single trait trio-PGS”), a model with all PGS traits included (“multi-trait trio PGS”) was also run for each of the six continuous measures. As a sensitivity analysis, these models were also run restricting the sample to only include one child per family to account for potential confounding from siblings sharing both genetic material and (likely) home environments. For these models the total variance and the variance explained due to direct and indirect effects by all the PGS combined were interpreted. The variance explained due to direct and indirect effects was calculated summing the partial *r*^2^ estimates of the PGS for mothers and fathers (indirect effects) and children (direct effects) in these models.

#### Software

All regression analyses and preparation of phenotypic data were run in R version 4.1.2 (R Core Team, 2021). Questionnaire data was ascertained using the MoBa phenotools (v3.0) R package ^[47]^. Trio-PGS regression models and estimation of clustered standard errors were conducted using the R packages stats (v4.1.2)^[48]^, MASS (v7.3-54)^[49]^, sandwich (v3.01)^[45,46,50]^, and lmtest (v0.9-38)^[51]^. Trio-GCTA models were run using the VCModels (v1.0.0)^[52]^ in version 1.6.2 of Julia^[53]^.

#### Preregistration and publicly available code

This study was preregistered. The preregistration is available at: https://doi.org/10.17605/OSF.IO/E2JQ3. Due to constraints on analytic possibilities resulting from the observed distributions of the data, we deviated from the preregistered analysis plan. An overview of the deviations and justifications for making them is presented in the Supplementary Note. Analytic code can be found at https://github.com/psychgen/within-fam-early-neurodev-moba.

## Results

For each of the measures of neurodevelopmental traits used in the main analyses, the maximum N available for analysis (i.e., with non-missing genotypic and phenotypic data) and distributional properties are shown in Table 2. Endorsement rates of the subdomains of motor (gross and fine motor) and language (expressive and receptive) development are shown in Supplementary Table 1.

**Table 2:** Descriptive information on the six continuous measures of early neurodevelopmental traits. Note-SD = standard deviation; Tr. = transformed scale.

| Scales | N | Min | Max | Mean | SD | Skew | Kurtosis | Tr. Skew | Tr. Kurtosis |
| --- | --- | --- | --- | --- | --- | --- | --- | --- | --- |
| Social & communication (SCQ) | 24599 | 0 | 22 | 2.23 | 1.73 | 1.74 | 7.2 | 0.58 | 1.07 |
| RRBI (SCQ) | 24558 | 0 | 12 | 3.77 | 2.49 | 0.63 | -0.09 |  |  |
| Attention (CBCL) | 24556 | 0 | 6 | 1.47 | 1.27 | 0.81 | 0.33 |  |  |
| Hyperactivity (CBCL) | 24550 | 0 | 6 | 1.96 | 1.37 | 0.42 | -0.11 |  |  |
| Motor (ASQ) | 24548 | 0 | 8 | 1.17 | 1.34 | 1.18 | 1.09 | 0.7 | -0.44 |
| Language (ASQ) | 24621 | 0 | 12 | 0.62 | 1.11 | 3.62 | 21.91 | 1.99 | 5.83 |

We tested the extent to which our analytic sample differed from the overall MoBa sample in two ways: by comparing individuals with and without genotyping information on the age 3 trait measures, and by comparing individuals with and without age 3 trait measures on the five PGS. The results are presented in Supplementary Tables 2-3. We found children with genotyping information available had significantly fewer reported neurodevelopmental difficulties in all areas, except motor development. Parents of and children with the age 3 data available had significantly lower PGS for ADHD and dyslexia (mothers and children only) and significantly higher PGS for educational attainment, cognitive ability, and autism (mothers only) then those missing the age 3 data.

### Trio-GCTA

The results of the Trio-GCTA model fitting are presented in Table 3. We find evidence for direct and, in some cases, indirect latent genome-wide genetic effects on early neurodevelopmental traits. A model with uncorrelated direct *and* indirect effects was the best fitting model using AIC criterion for attention, hyperactivity, and RRBI traits. For early language, motor, and social communication traits, the simpler model including only direct effects was favored.

**Table 3:**
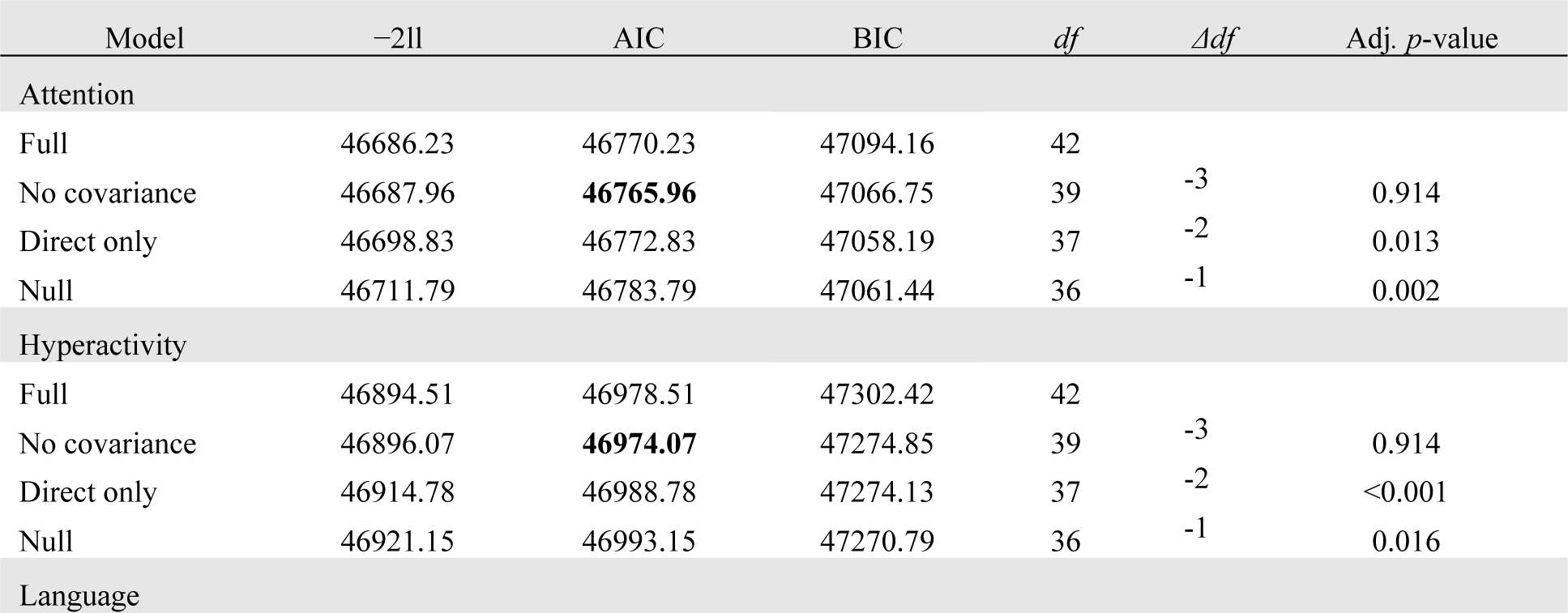
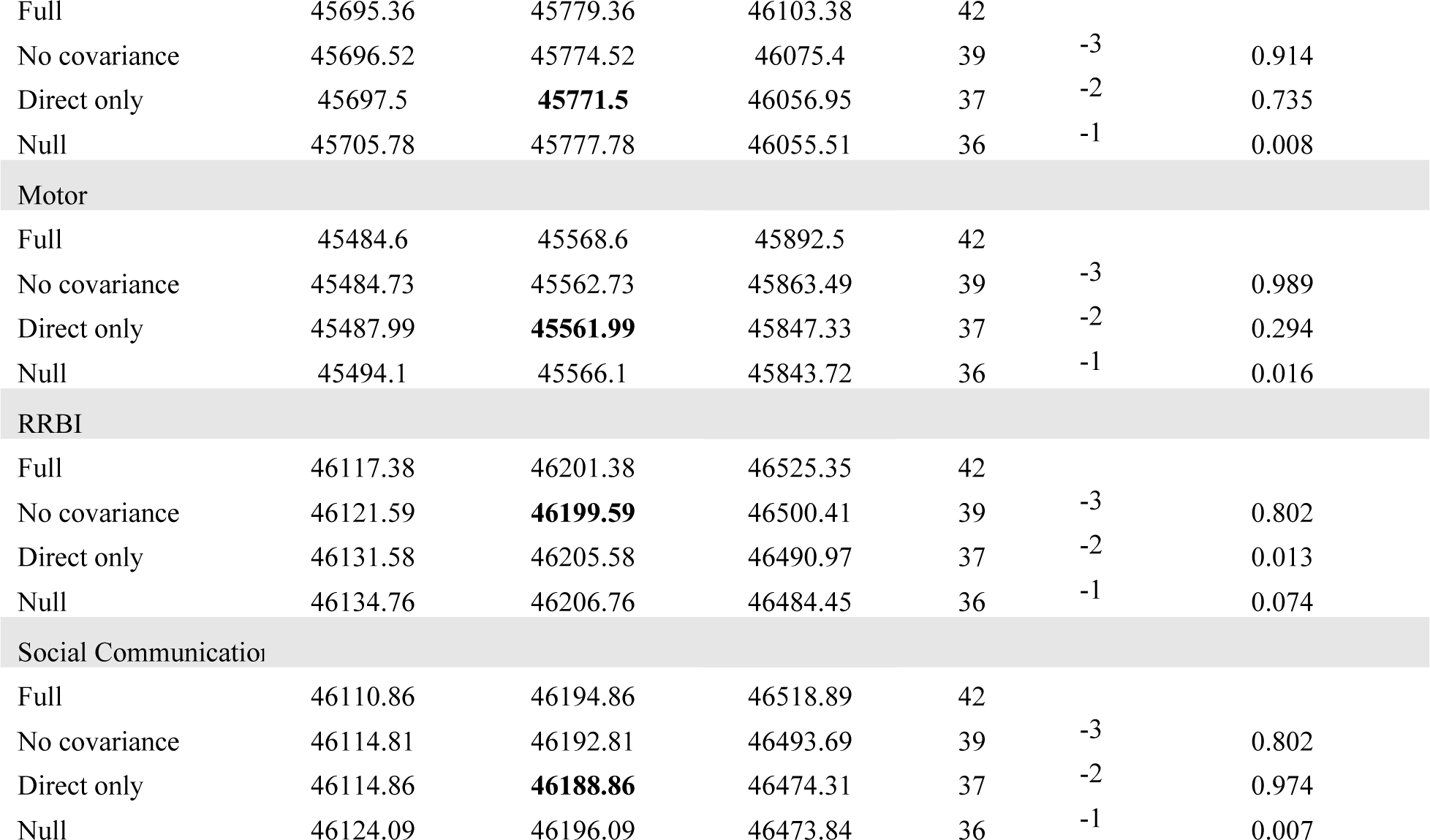
Akaike’s Information Criteria (AIC), Bayesian Information Criteria (BIC), and FDR corrected p-values for the likelihood ratio test presented for the four models run for each continuous neurodevelopmental outcome in the Trio-GCTA models. AIC bolded for best fitting model. −2ll = −2 log likelihood; df = degrees of freedom.

| Model | -2ll | AIC | BIC | *df* | $\Delta df$ | Adj. *p*-value |
| --- | --- | --- | --- | --- | --- | --- |
| Attention |  |  |  |  |  |  |
| Full | 46686.23 | 46770.23 | 47094.16 | 42 |  |  |
| No covariance | 46687.96 | **46765.96** | 47066.75 | 39 | -3 | 0.914 |
| Direct only | 46698.83 | 46772.83 | 47058.19 | 37 | -2 | 0.013 |
| Null | 46711.79 | 46783.79 | 47061.44 | 36 | -1 | 0.002 |
| Hyperactivity |  |  |  |  |  |  |
| Full | 46894.51 | 46978.51 | 47302.42 | 42 |  |  |
| No covariance | 46896.07 | **46974.07** | 47274.85 | 39 | -3 | 0.914 |
| Direct only | 46914.78 | 46988.78 | 47274.13 | 37 | -2 | <0.001 |
| Null | 46921.15 | 46993.15 | 47270.79 | 36 | -1 | 0.016 |
| Language |  |  |  |  |  |  |
| Full | 45695.36 | 45779.36 | 46103.38 | 42 |  |  |
| No covariance | 45696.52 | 45774.52 | 46075.4 | 39 | -3 | 0.914 |
| Direct only | 45697.5 | **45771.5** | 46056.95 | 37 | -2 | 0.735 |
| Null | 45705.78 | 45777.78 | 46055.51 | 36 | -1 | 0.008 |
| Motor |  |  |  |  |  |  |
| Full | 45484.6 | 45568.6 | 45892.5 | 42 |  |  |
| No covariance | 45484.73 | 45562.73 | 45863.49 | 39 | -3 | 0.989 |
| Direct only | 45487.99 | **45561.99** | 45847.33 | 37 | -2 | 0.294 |
| Null | 45494.1 | 45566.1 | 45843.72 | 36 | -1 | 0.016 |
| RRBI |  |  |  |  |  |  |
| Full | 46117.38 | 46201.38 | 46525.35 | 42 |  |  |
| No covariance | 46121.59 | **46199.59** | 46500.41 | 39 | -3 | 0.802 |
| Direct only | 46131.58 | 46205.58 | 46490.97 | 37 | -2 | 0.013 |
| Null | 46134.76 | 46206.76 | 46484.45 | 36 | -1 | 0.074 |
| Social Communication |  |  |  |  |  |  |
| Full | 46110.86 | 46194.86 | 46518.89 | 42 |  |  |
| No covariance | 46114.81 | 46192.81 | 46493.69 | 39 | -3 | 0.802 |
| Direct only | 46114.86 | **46188.86** | 46474.31 | 37 | -2 | 0.974 |
| Null | 46124.09 | 46196.09 | 46473.84 | 36 | -1 | 0.007 |

The estimated variance components for best-fitting and alternative Trio-GCTA models including genetic effects are shown in Figure 1. Here, indirect effects include the variance explained by the genotypes of the mother and father, while direct effects are the variance explained by the genotype of the child. In the full models, negative estimates are possible only in the case of covariances between direct and indirect effects, and where estimated these can increase the variance explained in early neurodevelopmental traits by allowing the same child and parental genetic variants to have partially opposite effects (left-most bar of each panel). However, as described above, all model fit statistics were in favor of simpler models not including these parameters. The estimates from the best fitting model for each outcome are indicated with an asterisk in Figure 1.

**Figure1:**
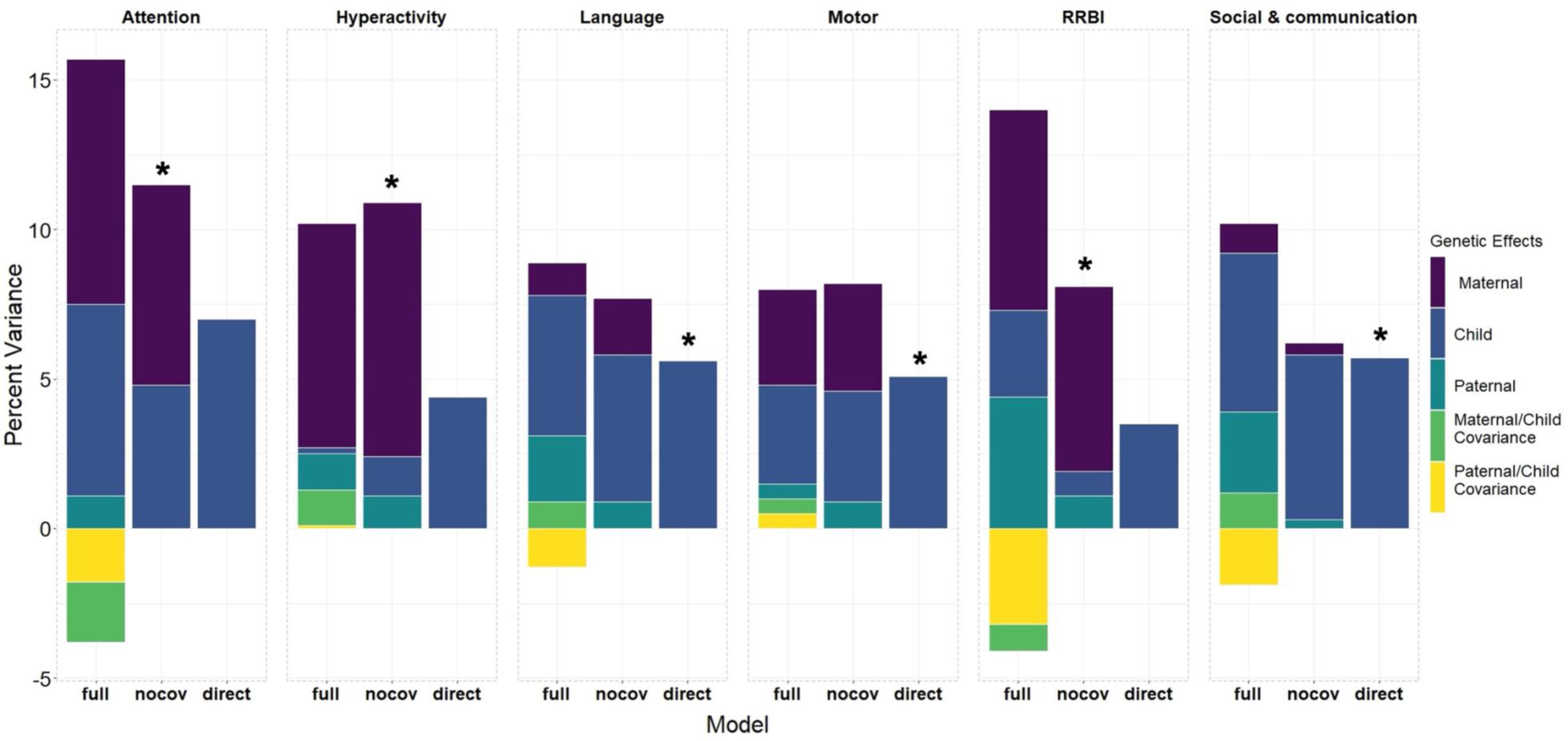
Variance component estimates for the full model estimating all parameters, the model not including covariances (nocov), and the direct effects only model. The best fitting model based on AIC is indicated with a “*”. Indirect effects include the variance explained by the genotypes of the mother and father while direct effects are the variance explained by the genotype of the child. In the full models, the covariance between the mother and child’s genetic effects and father and child’s genetic effects is estimated. This represents the extent to which the same genetic variation contributes to maternal/paternal direct and indirect genetic effects. When this covariance is positive this will increase the total variability genetic effects explained in the outcome. However, when the covariance between direct and indirect effects is negative, the true proportion of variance explained in the outcome by direct and indirect genetic effects is reduced as the effects cancel each other out.

In the models with the lowest AIC, direct genetic effects accounted for 4.8% [*SE* = 2.4%] of the variance in early attentional traits, 1.3% [*SE* = 2%] in hyperactivity, 0.8% [*SE* = 2.3%] in RRBI, 5.6% [*SE* = 2%] in language development, 5.1% [*SE* = 2.1%] in motor development, and 5.7% [*SE* = 2%] in social communication traits. Surprisingly, indirect genetic effects, and particularly maternal genetic effects, explained more variance relative to direct effects for hyperactivity (maternal indirect: 8.5% [*SE* = 2.1%], paternal indirect: 1.1% [*SE* = 2.1%]) and RRBI (maternal indirect: 6.2% [*SE* = 2.1%], paternal indirect: 1.1% [*SE* = 2.1%]). For attention, the magnitude of maternal indirect genetic effects (6.7% [*SE* = 2.7%]) was similar to the estimate for direct genetic effects. Paternal indirect effects did not explain any variability in early attentional traits. For language, motor, social communication, models without indirect effects were preferred. Variance component estimates for all models are reported in Supplementary table 4.

### Single-trait trio-PGS regression models

In the trio-PGS analyses, we found that direct genetic effects on early neurodevelopmental traits are currently not well captured by PGS of neurodevelopmental conditions. The results for these models run with the ADHD and autism PGS are shown in Figure 2. In the main analyses, the only direct genetic effects surviving correction for multiple testing was the ADHD polygenic score on attentional difficulties (*β* = 0.04 [0.02 – 0.05]) and for the autism polygenic score on language development (*β* = 0.02 [0.01 – 0.04]) and attentional difficulties (*β* = −0.02 [−0.04 – −0.01]). We did not find any significant effects for the dyslexia polygenic score in the main analysis (Supplementary Figure 1). In the sensitivity analyses including subscales of language and motor development, the only direct effect surviving multiple testing was of the dyslexia PGS decreasing the likelihood of any reported *gross* motor difficulties (Supplementary Figure 2).

Indirect effects surviving multiple testing of these PGS were found exclusively in fathers. The fathers’ polygenic score for ADHD was associated with attention (*β* = 0.02 [0.005 – 0.03]) and motor development (*β* = −0.02 [−0.04 – −0.01]) while the autism (*β* = −0.02 [−0.04 – −0.01]) and dyslexia (*β* = 0.02 [0.004 – 0.03]) polygenic score were associated with language development. These effects were still present when modeling the subdomains of motor and language development, being associated with fine motor skills (ADHD) and expressive language (autism and dyslexia).

**Figure 2:**
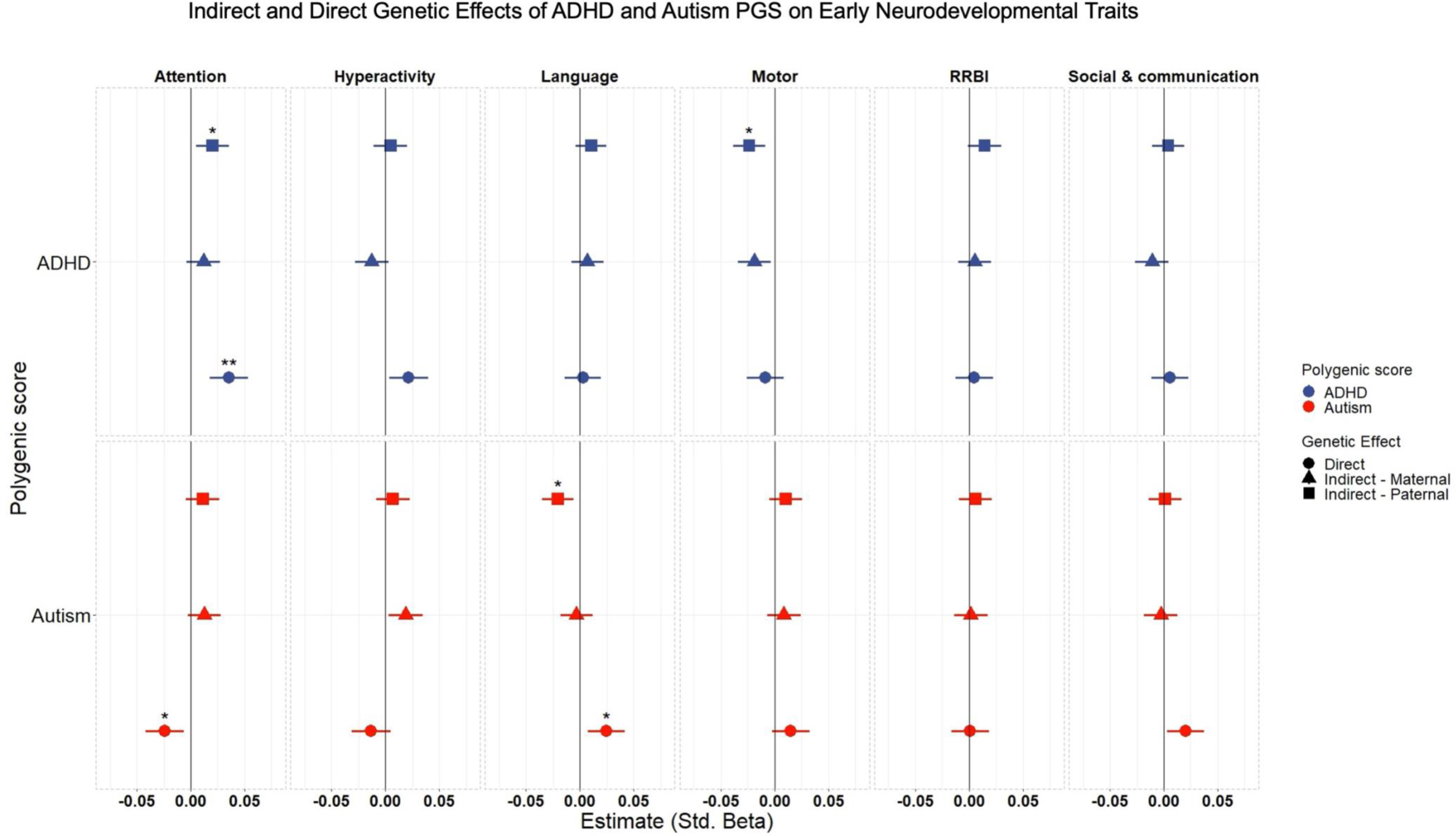
Standardized beta estimates for ADHD and Autism PGS on the six measures of early neurodevelopmental traits. 95% confidence intervals are shown. “*”, “**”, “***” denote adjusted p-values <0.05, <0.01, and <0.001 after multiple testing correction. All results presented are the PGS effect adjusting for the effect of the PGS for the other members of the trio.

Figure 3 shows the results for single-trait trio-PGS models of early neurodevelopmental traits regressed on cognitive ability and educational attainment PGS. Educational attainment and cognitive ability were seen to contribute to both direct and indirect effects on the neurodevelopmental traits. Direct effects surviving multiple testing were observed for both educational attainment (*β* = −0.05 [−0.06 – −0.03]) and cognitive ability (*β* = −0.04 [−0.06 – −0.03]) on attention, and for cognitive ability only on language (*β* = −0.03 [−0.05 – −0.01]) and hyperactivity (*β* = −0.03 [−0.04 – −0.01]).

Either one or both of the educational attainment and cognitive ability PGS indicated indirect effects (either maternal, paternal, or both) on all domains of neurodevelopment. For the motor domain, we found that both maternal and paternal PGS for educational attainment and cognitive ability were significantly associated with the measured phenotype. When effects were modeled at the level of fine and gross motor sub-domains, we observed that the indirect effects on motor development were exclusive to fine motor skills (Supplementary Figure 2). In addition, a significant positive direct genetic effect of educational attainment on gross motor skill difficulties was present that was not found for the total motor score. Model parameters for the single PGS-trait models are reported in Supplementary Table 5.

**Figure 3:**
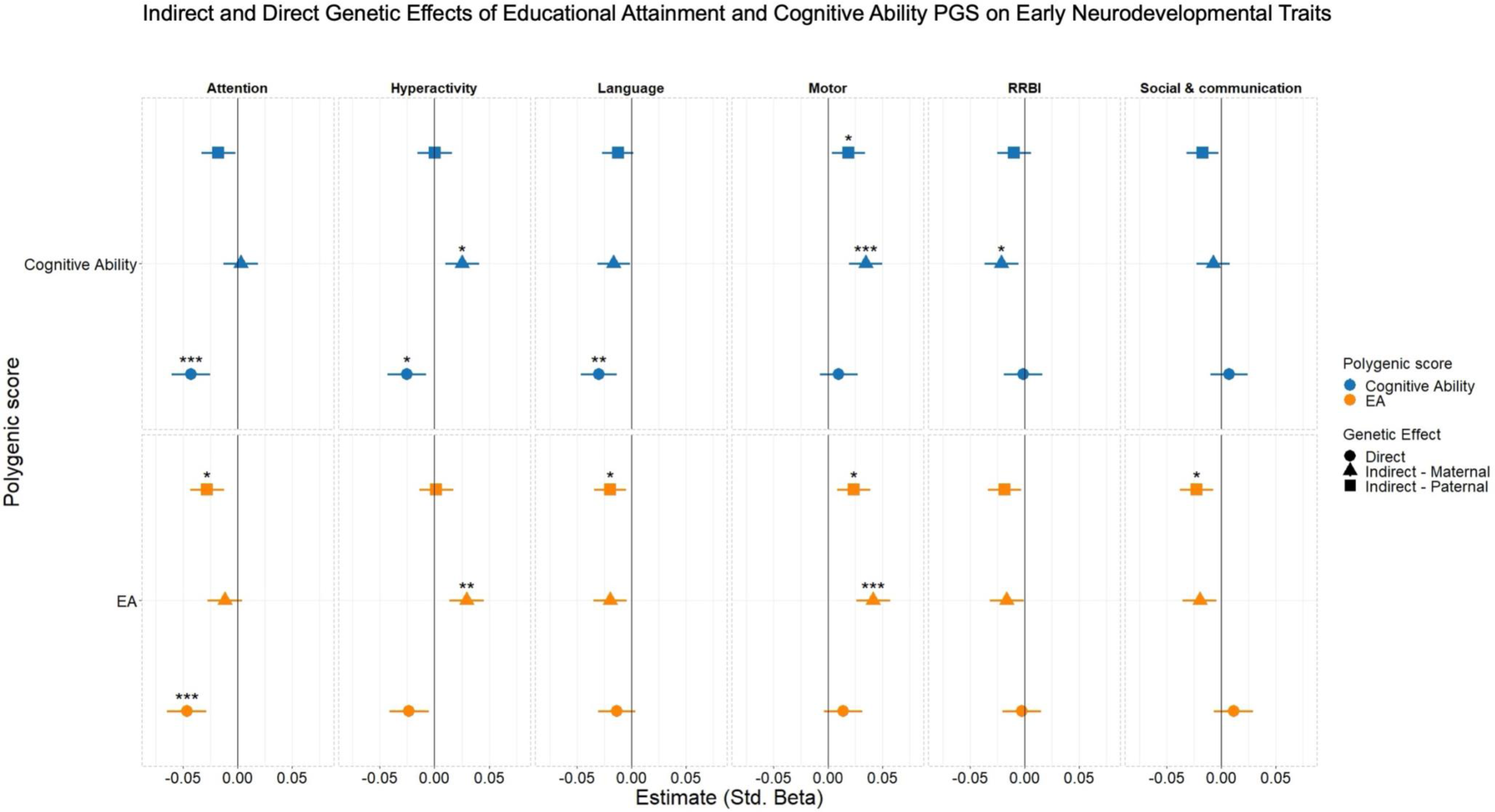
Standardized beta estimates for cognitive ability and educational attainment (EA) PGS on the six measures of early neurodevelopmental traits. 95% confidence intervals are shown. “*”, “**”, “***” denote adjusted p-values <0.05, <0.01, and <0.001 after multiple testing correction. All results presented are the PGS effect adjusting for the effect of the PGS for the other members of the trio.

### Multi-trait trio-PGS regression models

PGS for autism, ADHD, dyslexia, educational attainment, and cognitive ability – included for all members of a family trio – jointly explained very little variance in early neurodevelopmental traits. Together, these scores’ direct effects only explain 0 – 0.13% of the variance and indirect effects explain 0.04 – 0.14% of variance in the neurodevelopmental traits (Supplementary Table 6).

Sensitivity analyses removing siblings had no effects on the estimates (Supplementary Table 6). In the multi-trait models, direct genetic effects explain more variance than indirect genetic effects for attention while the opposite was true for hyperactivity, motor, RRBI, and social communication traits, although no formal comparison was made. Direct *vs.* indirect effect contributions were similar for early language development (Supplementary Table 6).

## Discussion

In this pre-registered study, we estimated the relative contributions of direct and indirect genetic effects for a range of maternally reported behavioral traits linked to neurodevelopmental conditions in 3-year-old children sampled from the Norwegian population. We did this using two complementary methodological approaches. Firstly, by focusing on partitioning *overall* genetic influence on early neurodevelopmental traits into direct and indirect effects and secondly, seeking evidence for the sources of those effects in specific, pre-established genetic liabilities. With the first approach, we found evidence for both direct and indirect latent genetic effects on attention, hyperactivity, and restrictive and repetitive behaviors among 3-year-old children. Whilst for language, motor, and social communication traits at the same age we found evidence for only direct genetic effects. Genetic liability for ADHD, autism, cognitive ability, and educational attainment provided specific sources of direct genetic effects for attention. Likewise, genetic liability for autism and cognitive ability were a source of direct genetic effects on language development, as well as cognitive ability on hyperactivity. We found that genetic liability for cognitive ability and/or educational attainment were sources of maternal and/or paternal indirect effects for all neurodevelopmental outcomes while indirect effects attributable to genetic liability for autism, ADHD, and dyslexia were more specific and only found in fathers.

Direct genetic effects, usually implicitly assumed to be the predominant component of heritability estimates, were indeed robustly evident for almost all neurodevelopmental traits in our Trio-GCTA models. While this was not replicated across all traits in the trio-PGS analyses, this should do little to undermine our confidence that these effects exist given the tiny fraction of the SNP heritability (and overall variance) in outcomes that current PGS explain. Instead, we conclude only that the direct effects observed at the genome-wide level are not well captured by the PGS included in the analyses. Including additional PGS for other neurodevelopmental conditions and related traits as well as future versions of included scores based on more progressed GWAS would likely eventually capture a larger proportion of these effects. Differences between the underlying GWAS traits, populations, and sample sizes, the presence and magnitude of effects across different PGS are not easily comparable so little should be read into comparison of genetic liabilities for neurodevelopmental conditions *versus* educational attainment and cognitive ability. However, what *is* noteworthy here is that educational attainment – and, to a lesser extent, cognition – show substantially attenuated heritabilities in within-family GWAS ^[21]^, suggesting these scores in particular are likely to include indirect genetic effects or more broad structural confounding. Here, consistent with the within-family GWAS results, we observe that both direct and indirect effects on early neurodevelopmental traits are captured in these scores.

In the Trio-GCTA analyses, for three traits (attention, hyperactivity, RRBI) we found evidence for indirect genetic effects of surprisingly large relative magnitude: either exceeding or equaling direct genetic effects in the same models in all cases. This sits in contrast to results from the same sample, using the same and similar methodologies to investigate 8-year ADHD traits, where direct effect estimates were invariably at least twice as large as indirect equivalents.^[25,26]^ It should be noted that, in both cases, effects in question are quite imprecisely estimated. Nonetheless, such a developmental shift, if confirmed with formal comparisons, would be consistent with a developmental model of decreasing passive gene-environment correlation over time, as parental influences wane ^[54]^. However, elevated indirect effects – in particular, maternal indirect effects – at early time points are also consistent with a greater influence of maternal (genetically-influenced) rater bias when children are younger. Additionally, potential maternal rater bias may give some insight into the somewhat counterintuitive direction of the effect observed in several of the polygenic score analyses (i.e. we find higher maternal (and not paternal) polygenic liability to educational attainment is positively associated with higher reported hyperactivity traits). However, further work is needed to conclusively determine any of the mechanisms underpinning these results. More generally, we observe relatively low proportions of the variance explained by SNPs across all traits in the Trio-GCTA models – here low reliabilities of the trait measures and limited endorsement of some traits may have influenced results.

For some traits we see concordance in the findings between the two modeling approaches. For example, direct effects explain 4.8% of the variance in attention scores at age 3, and this is partly attributable to genetic liabilities for ADHD, autism, educational attainment, and cognitive ability. Similarly, indirect effects explaining 8.5% of the variance in hyperactivity can, according to the trio-PGS results, be traced in part to maternal genetic liability for educational attainment. However, we also observe seemingly inconsistent results – for example, no overall indirect genetic effects were detected for motor development in the best fitting Trio-GCTA model, yet maternal genetic liability for cognitive ability and educational attainment and paternal genetic liability for ADHD are identified as significant sources of such effects in the trio-PGS. Here, a key point should be noted to resolve this apparent inconsistency – and, in turn, dampen enthusiasm towards the consistencies outlined above. This being the scale at which PGS analyses currently operate, as PGS explain a tiny fraction of variance both collectively and on their own. In partitioning variance into latent genome-wide genetic effects, the significant effects captured by the PGS are likely too small to influence the “overall picture” assessed by the Trio-GCTA models. Put another way, the model-fitting used in Trio-GCTA – even with samples of this size – is something of a blunt instrument, often allowing simpler models to be preferred based on sometimes marginal differences in fit. Nonetheless, it should be emphasized that these methods rely on some different assumptions about the data, and the processes underlying genetic effects. We propose here that their application in parallel allows for a more complete discussion of direct and indirect effects – at both the macro and micro scale.

## Limitations

There are some limitations that should be considered in the interpretation of these results. Firstly, although the polygenic score direct effect estimates found in these analyses are *more* robust to bias from indirect genetic effects, assortative mating, and population stratification than estimates *not* adjusting for parental scores, they are still observational estimates still best interpreted as associations rather than causal effects *per se*. This may be particularly relevant where the mechanisms for confounding are dynastic, such as assortative mating in generations prior to the parent generation included here, or structural at the GWAS stage, such as residual population stratification and selection into the population based GWAS used to construct the PGS ^[55]^. The Trio-GCTA method, which estimates direct effects via differences in genome-wide relatedness rather than identifying specific causal loci may be more robust to these factors. ^[56]^ Nonetheless, the key consideration remains that estimates of indirect effects in both trio-PGS and Trio-GCTA can be influenced by both the assumed genetic nurture effect but also by a range of different biases, including assortative mating, residual population stratification, and unmeasured effects of other related individuals including siblings and broader multi-generational (“dynastic”) effects ^[25,55,57]^. Also important to the interpretation are potential biases arising from both selection and attrition in MoBa, particularly in relation to factors relevant to the traits in the present study ^[58]^ – such as the overrepresentation of families with higher educational levels and socioeconomic backgrounds in MoBa. ^[59]^ Additionally, we observed that families of children with neurodevelopmental conditions were lost to follow up at higher rates than the rest of the cohort. Finally, these analyses were limited to those of European ancestry, limiting the generalizability of these results.

## Conclusions

Understanding the genetic contributions underlying early neurodevelopmental traits in a within-family paradigm can give insight into the various pathways of genetic transmission involved in the development of neurodevelopmental conditions. We find direct genetic effects contribute to all early neurodevelopment traits in 3-year-old children, with differing levels of importance across the areas of neurodevelopment. We also find evidence that indirect genetic effects may be particularly relevant to explaining variability in early maternally reported measures of attentional traits, hyperactivity, and restricted and repetitive behaviors and interests. We find evidence of both direct and indirect effects, albeit small, being captured by PGS for neurodevelopmental conditions and related traits. Overall, we find direct and indirect genetic effects influence neurodevelopmental traits at age 3 to varying degrees, highlighting the importance of within-family approaches for disentangling genetic processes that influence the traits and their relation to neurodevelopmental conditions.

## Data Availability

The consent given by the participants does not allow for storage of data on an individual level in repositories. Researchers can apply for access to data for replication purposes via MoBa, in line with MoBa data access policies.

## Acknowledgements

The Norwegian Mother, Father and Child Cohort Study is supported by the Norwegian Ministry of Health and Care Services and the Ministry of Education and Research. We are grateful to all the participating families in Norway who take part in this on-going cohort study. We thank the Norwegian Institute of Public Health (NIPH) for generating high-quality genomic data. This research is part of the HARVEST collaboration, supported by the Research Council of Norway (#229624). We also thank the NORMENT Centre for providing genotype data, funded by the Research Council of Norway (#223273), South East Norway Health Authorities and Stiftelsen Kristian Gerhard Jebsen, and in collaboration with deCODE Genetics. We further thank the Center for Diabetes Research, the University of Bergen for providing genotype data funded by the ERC AdG project SELECTionPREDISPOSED, Stiftelsen Kristian Gerhard Jebsen, Trond Mohn Foundation, the Research Council of Norway, the Novo Nordisk Foundation, the University of Bergen, and the Western Norway Health Authorities.

This work was performed on the TSD (Tjeneste for Sensitive Data) facilities, owned by the University of Oslo, operated and developed by the TSD service group at the University of Oslo, IT Department (USIT). Analyses were performed on resources provided by Sigma2 - the National Infrastructure for High-Performance Computing and Data Storage in Norway.

