## Supplementary Material for "Direct and Indirect Genetic Effects on Early Neurodevelopmental Traits"

### **Supplementary Note**

*Note on deviations from the pre-registration*

Due to constraints on analytic possibilities resulting from the observed distributions of the data, we were forced to deviate from the preregistered analysis plan. The original pre-registration can be found on the OSF repository here: <https://osf.io/zwk2a> and the updated document here: <https://osf.io/xjfw9>

An overview of the reasoning behind the deviations:

**Removal of bifactor model:**  The original version of the preregistration for this study included modeling general and domain-specific factors underlying early neurodevelopmental traits in an SEM framework, using the items from the scales as indicators of these factors. However, the initial investigation revealed that both the planned broad version (four domain-specific factors) and narrow version (eight domain-specific factors) of these bifactor models provided a poor fit to the data with substantial evidence for model misspecification. Exploratory factor analysis that included an expanded set of items from those included in this analysis found that a correlated factor model was a substantially better way of explaining covariance between early neurodevelopmental items, indicating a large amount of heterogeneity underlying these traits in the general population and little evidence for a shared general factor (Hegemann et al., 2023). Therefore, instead of proceeding with a misspecified model we chose to remove the bifactor model from the analysis. Because of the indication of many factors underlying these early traits in general populations we chose to move forward breaking the scales into subscales in accordance with our original “narrow” version of the specific factors. Allowing us to look at estimates of genetic effects with more specificity than using the “broad” version. Because of the distributions of the motor and language subscales, we further deviated from this. We used the “broad” version of the motor and language subscales in order to have a continuous measure of these domains for the trio-GCTA analyses and, for consistency, used these in addition to the “narrow” versions in the trio-PGS analyses.

**Use of observed scores over factor scores:** Because of the removal of the bifactor model there was no longer the need for estimating general and specific components underlying variance in early neurodevelopmental traits from the items so observed scores could be used instead.

**Removal of the general and specific measurement model analyses:** Due to the poor fit and evidence of misspecification of the bifactor model we concluded that it was not feasible to move forward with further models based on it.

**Changes to PGS:** The ADHD and dyslexia PGS were created with the more recent GWAS for both traits that had become available after the initial pre-registration. A PGS created from the UK biobank handgrip measure was dropped from the updated pre-registration, prior to running to the trio-PGS models. Originally the score was included under the logic that it was the best proxy available for motor development. However, under further consideration we concluded that the PGS analyses should be limited to PGS for neurodevelopmental conditions and traits that have substantial literature backing there associations to these conditions.

**Supplementary Methods**

Full instrument documentation on the MoBa questionnaires can be found elsewhere: <https://www.fhi.no/en/ch/studies/moba/for-forskere-artikler/questionnaires-from-moba/#36-months-after-birth>

The following instruments where used for the measures of early neurodevelopmental traits:

| Outcome | Instrument | Items (codes in MoBa questionnaire – see instrument documentation) |
| --- | --- | --- |
| Social & communication | SCQ | GG256, GG257, GG259, GG264, GG265, GG274-88, GG290-94 |
| RRBI | SCQ | GG258, GG260-3, GG266-71, GG273 |
| Attention | CBCL | GG314, GG315, GG332 |
| Hyperactivity | CBCL | GG316, GG320, GG327 |
| Motor | ASQ – Motor | GG222-5 |
| Language | ASQ – Communication | GG237-42 |
| Fine Motor | ASQ – Motor | GG224, GG225 |
| Gross Motor | ASQ – Motor | GG222, GG223 |
| Expressive Language | ASQ – Communication | GG239, GG241, GG242 |
| Receptive Language | ASQ – Communication | GG237, GG238, GG240 |

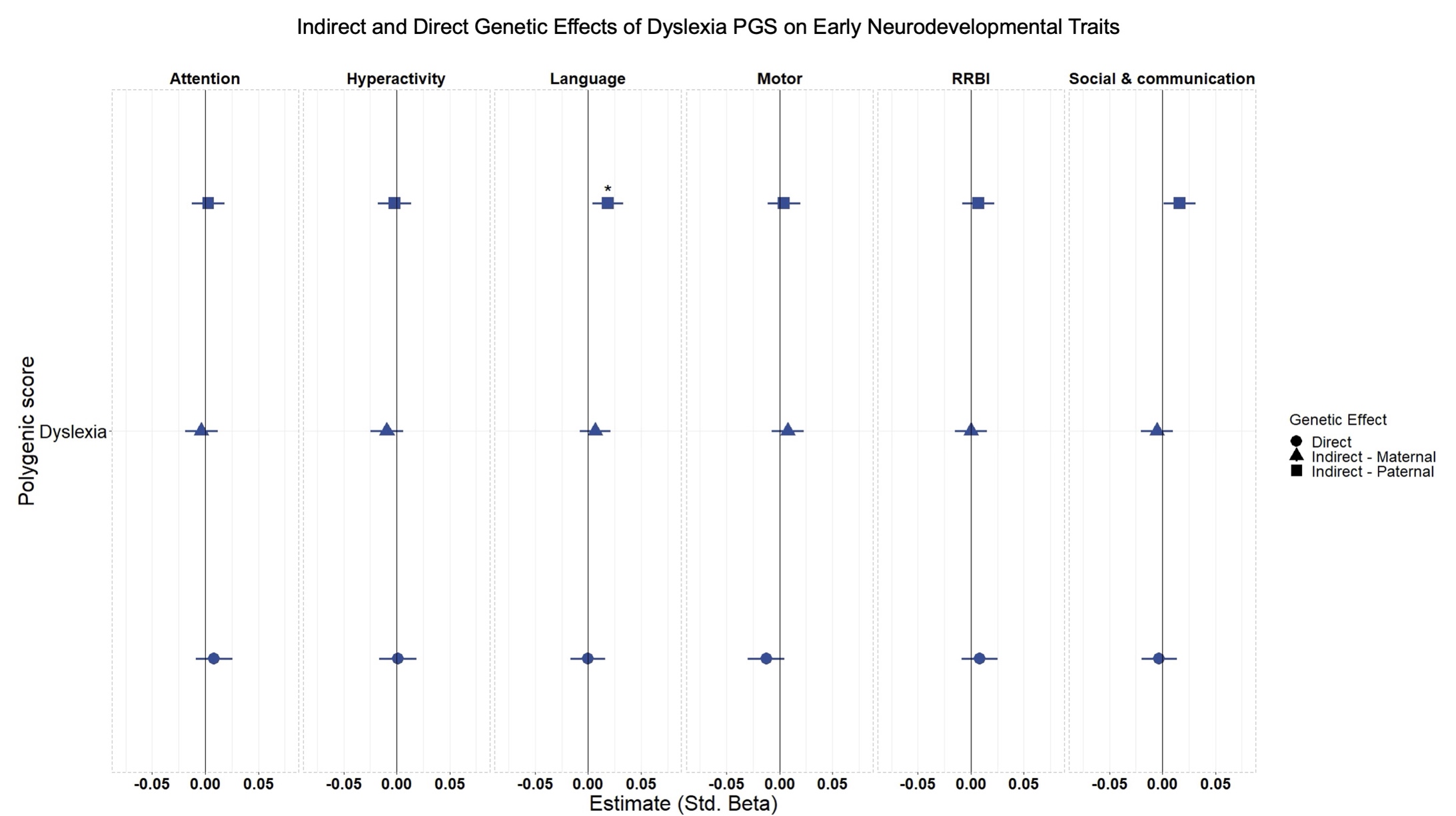

### **Supplementary Figures**

Figure S1: Standardized beta estimates for dyslexia PGS on the six measures of early neurodevelopmental traits. 95% confidence intervals are shown. “*”, “**”, “***” denote adjusted p-values <0.05, <0.01, and <0.001 after multiple testing correction. All results presented are the PGS effect adjusting for the effect of the PGS for the other members of the trio.

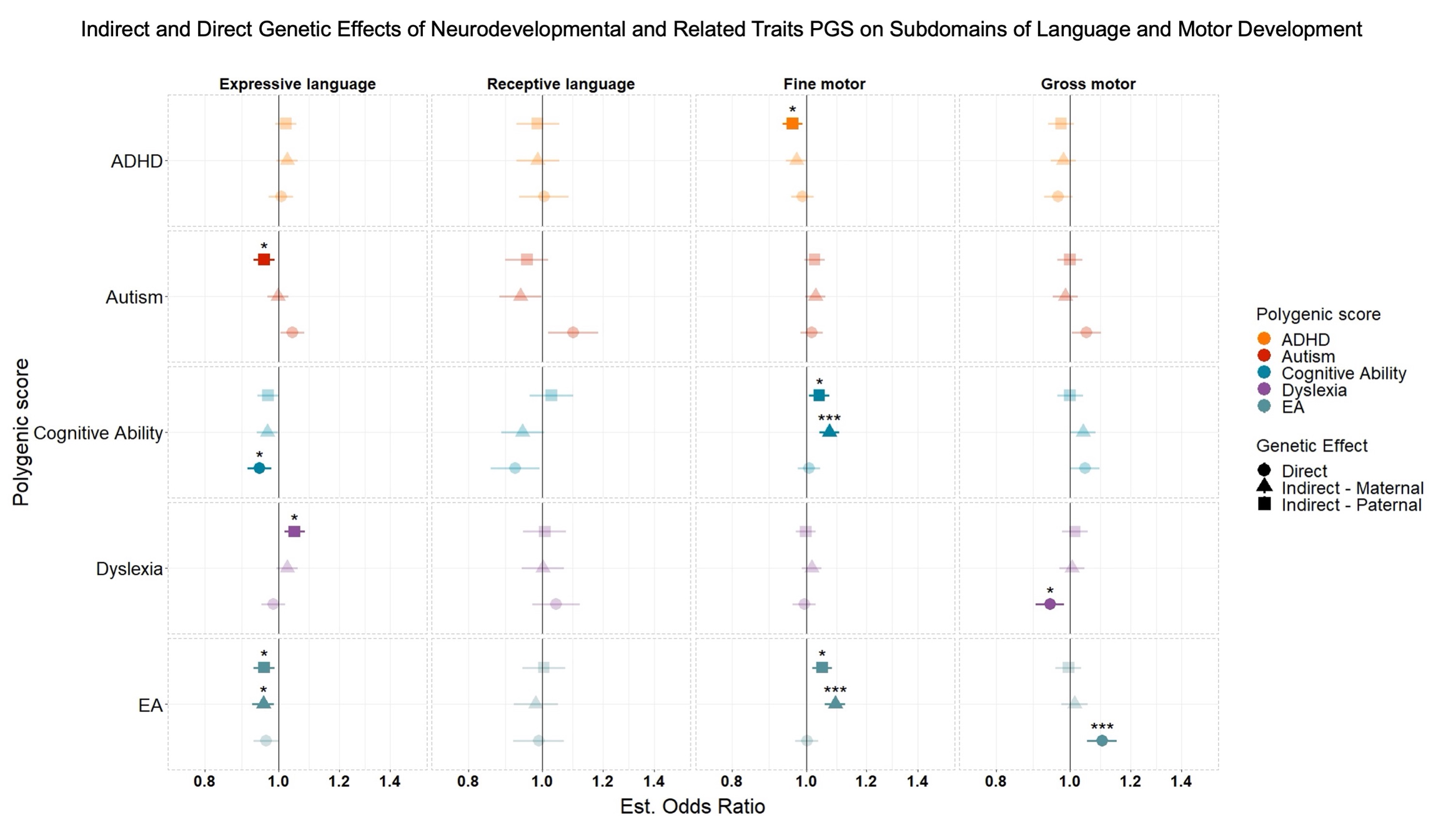

Figure S2: estimated log odds ratio for all PGS on reporting difficulties in the subdomains of language and motor difficulties. 95% confidence intervals are shown. “*”, “**”, “***” denote adjusted p-values <0.05, <0.01, and <0.001 after multiple testing correction. All results presented are the PGS effect adjusting for the effect of the PGS for the other members of the trio.
